# Factors associated with access to Insecticide-Treated Nets and use of house modification in Bagamoyo and Ulanga districts, Tanzania

**DOI:** 10.1101/2019.12.16.19015081

**Authors:** OG Odufuwa, A Ross, YP Mlacha, O Juma, S Mmbaga, DF Msellemu, SJ Moore

**Affiliations:** Ifakara Health Institute, Tanzania; Swiss Tropical and Public Health Institute, Basel, Switzerland; University of Basel, Basel, Switzerland

**Keywords:** Access, Insecticide Treated Nets, ITNs, House modification, Malaria, Demographic, Socioeconomic, Breeding sites, Tanzania

## Abstract

**Background:** Insecticide-treated Nets (ITNs) and house modifications are proven vector control tools, yet full coverage has frequently not been achieved. This study investigates factors associated with ITN access and use of house modification in Tanzania.

**Methods:** Baseline data from cross-sectional surveys was analysed from 7,240 households in Bagamoyo (60km north of Dar es Salaam) and 1,241 households in Ulanga (a remote rural area), from previous studies on spatial repellants and indoor residual spray evaluation. We estimated the associations between the outcomes: population access to ITNs, ITN per sleeping spaces, presence of window screens and closed eaves, and the covariates household size, age, gender, pregnancy, education, mosquito entry (doors and windows), house modification (window screens & closed eaves), mean distance to mosquito breeding sites and wealth using regression models.

**Results:** Population access to ITNs (households with one ITN per two potential people that stayed in the house the previous night of the survey) was 64% (56 - 71) and ITNs per sleeping spaces (households with enough ITNs to cover all sleeping spaces used the previous night of the survey) was 63% (54 - 72) in Bagamoyo, three years after the last Universal Coverage Campaigns (UCC). These findings are both lower than the 80% coverage target of the Tanzania National Malaria Strategic Plan (Tanzania NMSP). In Ulanga, population access to ITNs was 84% (78 - 91) and ITNs per sleeping spaces was 93% (89 - 97), one year after the last UCC. Household size was significantly associated with lower access to ITNs even shortly after UCC. House modification was common in both areas. In Bagamoyo, screened windows were more common than closed eaves (61% vs 14%) whereas in Ulanga more houses had closed eaves than window screens (55% vs 12%). Households in the poorest quintile were less likely to adopt window screening and closing eaves in Bagamoyo, this contrasts with Ulanga district where a greater proportion of poorer households blocked eaves with mud but were less likely to adopt screening.

**Conclusion:** Population access to ITN was substantially lower than the targets of the Tanzania NMSP after three years and lower among larger households after one year following ITN campaign. House modification was common in both areas, motivated by wealth. Improved access to ITNs and window screens through subsidy, especially among larger households, uptake of a combination of these two interventions may be maximized.

## BACKGROUND

Insecticide-Treated Nets (ITNs) have made the greatest contribution to the reduction in malaria burden in sub-Saharan Africa [1], through both individual and especially community effects that reduce the proportion of mosquitoes living long enough to transmit malaria [2]. ITNs work best when applied at high (>80%) coverage [2]. Hence, ITNs are recommended by the World Health Organization (WHO) to be universally and continuously distributed to all people at risk of malaria [3]. Multiple ITN delivery strategies and campaigns have been implemented across sub-Saharan Africa, either targeting specific malaria risk populations (children under 5 years of age and pregnant women), or targeting the entire population using health facilities, antenatal clinics (ANC), schools, markets, door-to-door and other methods [4]–[7]. Despite more than a decade of ITN campaigns, 60% of the population in malaria endemic areas, including Tanzania still do not have access to an ITN [8], which means that far greater malaria control could be attained with improvement in access of this highly efficacious intervention.

House modification using window screens or blocking eaves (Figure 1) which serve as barriers against mosquito entry has been shown to be associated with lower malaria infection [9] and a control tool for other vector borne diseases [10]. However, house modification is not currently delivered operationally as a vector control tool, being generally considered too costly or difficult to implement [11]. Perhaps, because of high installation cost [12], there is limited support from government or other organisations [13] to promote house modification as a public health tool even though it provides protection from mosquito vectors when people are indoors but outside of sleeping hours (i.e. when people are not protected by ITNs) [14], and thereby can prevent malaria and other vector-borne diseases such as dengue. There is evidence of houses with window screens and closed eaves among the urban Tanzanian population due to their effectiveness in improving house ventilation and preventing mosquito entry and nuisance [15], [16].

**Figure 1:**
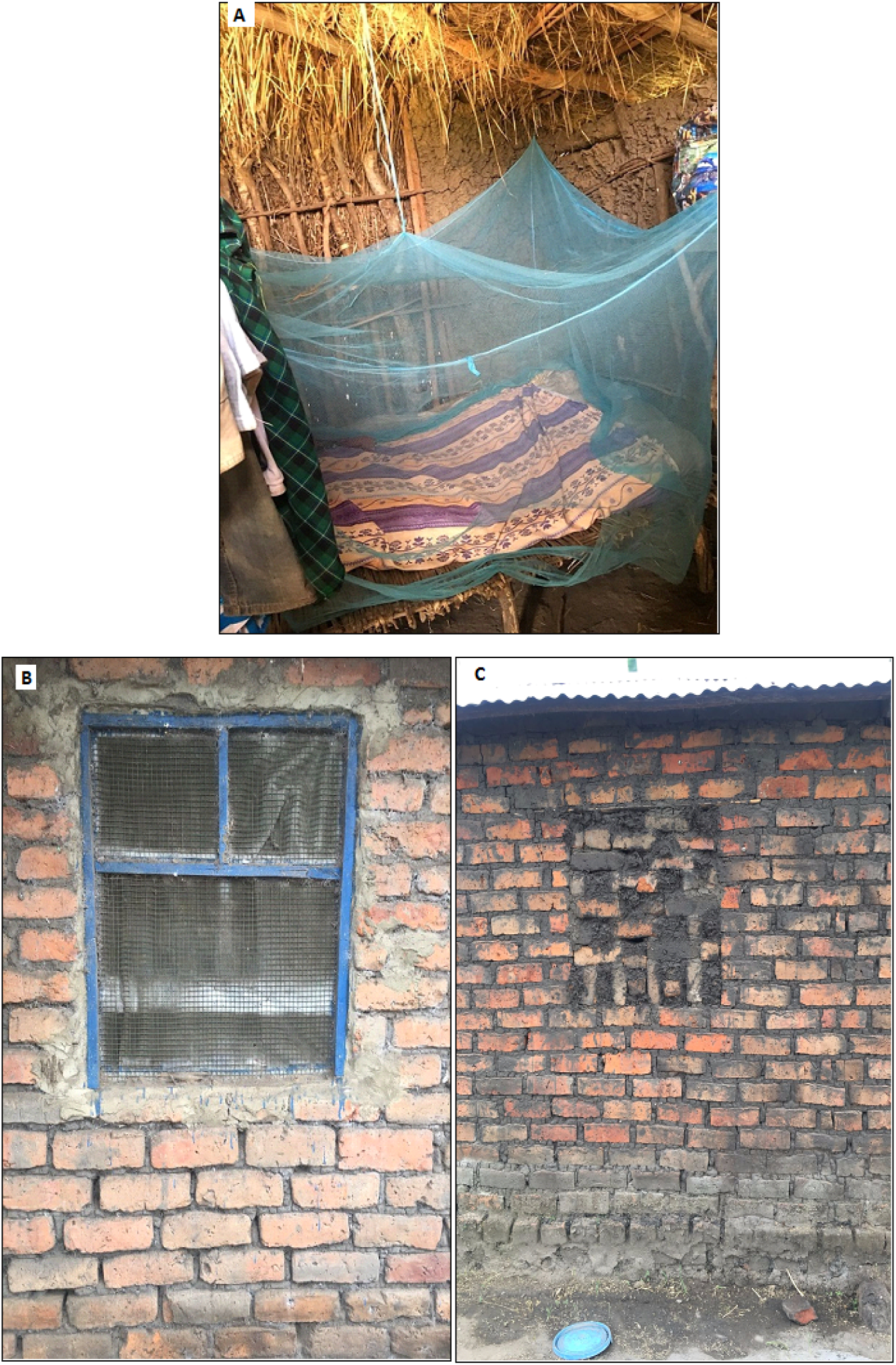
A. An ITN hanging in a rural Tanzanian home B. Screened window C. Eaves and ventilation bricks closed with mud to prevent mosquito entry.

This paper reports the drivers of access to ITNs and the use of house modification between two rural populations: one far from and one close to an urban centre, as it is known that access to economic centers affects the economic, education, and health status of populations [17]. As the NMCP in Tanzania is now stratifying vector control delivery systems (Tanzania ITN Plan 2016-2020, unpublished) in line with WHO policy [18]. This work is aimed to provide useful information to policy makers in maximising access to and use of these existing malaria control tools.

## METHODS

### Study area and demographic characteristics

The study was conducted in Bagamoyo and Ulanga districts (Figure 2). Bagamoyo district is located on the east coast of Tanzania, approximately 60 km north of Dar es Salaam, the economic hub of the country [19] and Ulanga district is located in rural south east Tanzania, 300km from the regional city of Morogoro and 500km from Dar es Salaam [20]. In Bagamoyo and Ulanga, the average household size was 4.4 and 4.9 persons per household, respectively in the 2012 Tanzania National Census [21]. Adult literacy in Bagamoyo was 58% and 66% in Ulanga [22]. The average rainfall and temperature are 1200 to 2100 mm per year and approximately 28°C in both districts, with slightly higher average rainfall per year and temperature in Bagamoyo due to its coastal location. Above 70% of the residents in both districts own pieces of land and majority of them engage in subsistence farming [19], [23]. The pattern of climatic conditions and land use are contributing factors toward the persistence of the perennial breeding sites for mosquitoes and other diseases vectors [24].

**Figure 2:**
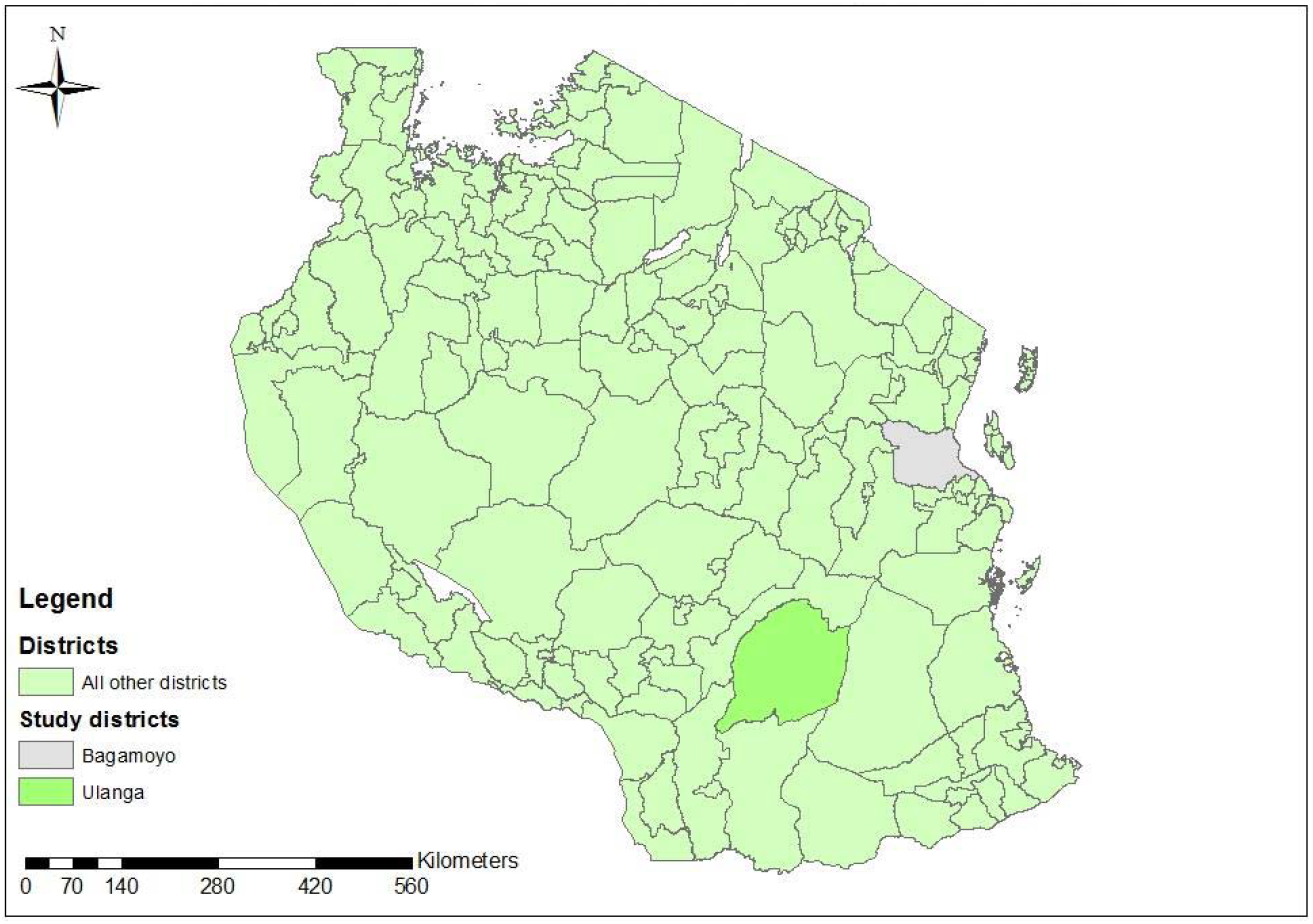
Geographical location of Bagamoyo and Ulanga districts in Tanzania.

### Study Design

The study is a secondary analysis of two baseline cross-sectional surveys from a trial on spatial repellents in Bagamoyo [25], and an insecticide residual spray (IRS) evaluation in Ulanga [26]. The surveys were conducted randomly in nine villages in Bagamoyo district from the year 2014 to 2015 and eight villages in Ulanga district from the year 2016 to 2017.

### Household survey data collection

Data were collected using a structured questionnaire upon written informed consent from an adult household member. The questionnaire surveyors collected information on ITN coverage indicators: 1) households with one ITN per sleeping space (proportion of households with enough ITN for all sleeping spaces) [27] and 2) proportion of the population with at least 1 ITN for every 2 people that slept in the household on average [28].

Information was collected on demographic and socio-economic characteristics (age, sex, household size, house structure, livestock ownership, asset ownership, source of light, occupation, level of education, and presence of pregnant occupants), household geographical coordinates (longitude, latitude and elevation) and house structure (wall materials, roof materials, ceiling materials, opened or closed eaves, floor materials, number of windows, if windows were screened, and number of outside doors were noted based on visual observation by the interviewers). The surveys were delivered using similar questionnaires in the two sites except that in Ulanga, data on age, sex and educational status of heads of households and the geographical coordinates were not collected.

### Data Management and Analysis

The paper questionnaires were double-entered using EpiData software [29] and analysis was carried out using STATA 14.2 [30].

Socioeconomic status of each household was derived from a weighted score using principal component analysis (PCA), categorized into quintiles: poorest, poor, middle, high and highest [31]. The binary outcomes of presence of window screens and presence of closed eaves were analyzed using a logistic regression model, while the count outcomes, ITN access and ITN per sleeping spaces were analysed using Poisson regression model. The associations between the outcomes and the covariates, household size, age, sex, pregnancy, education, mosquito entry (doors and windows), house modification (window screens and closed eaves), mean distance to breeding site and socioeconomic quintiles were estimated. Village was included in the regression model as a random effect to account for clustering. The age of the heads of households was categorized into three levels: young adults “18-24”, adult “25-49” and old adults “50-99”.

All covariates and interaction between education and wealth status were considered in the analysis based on findings from past literature [32]–[37]. Indicator of ITN access and enough ITN for sleeping spaces for this study were defined in accordance with the household surveys indicators for malaria control [38] (Figure 3).

**Figure 3:**
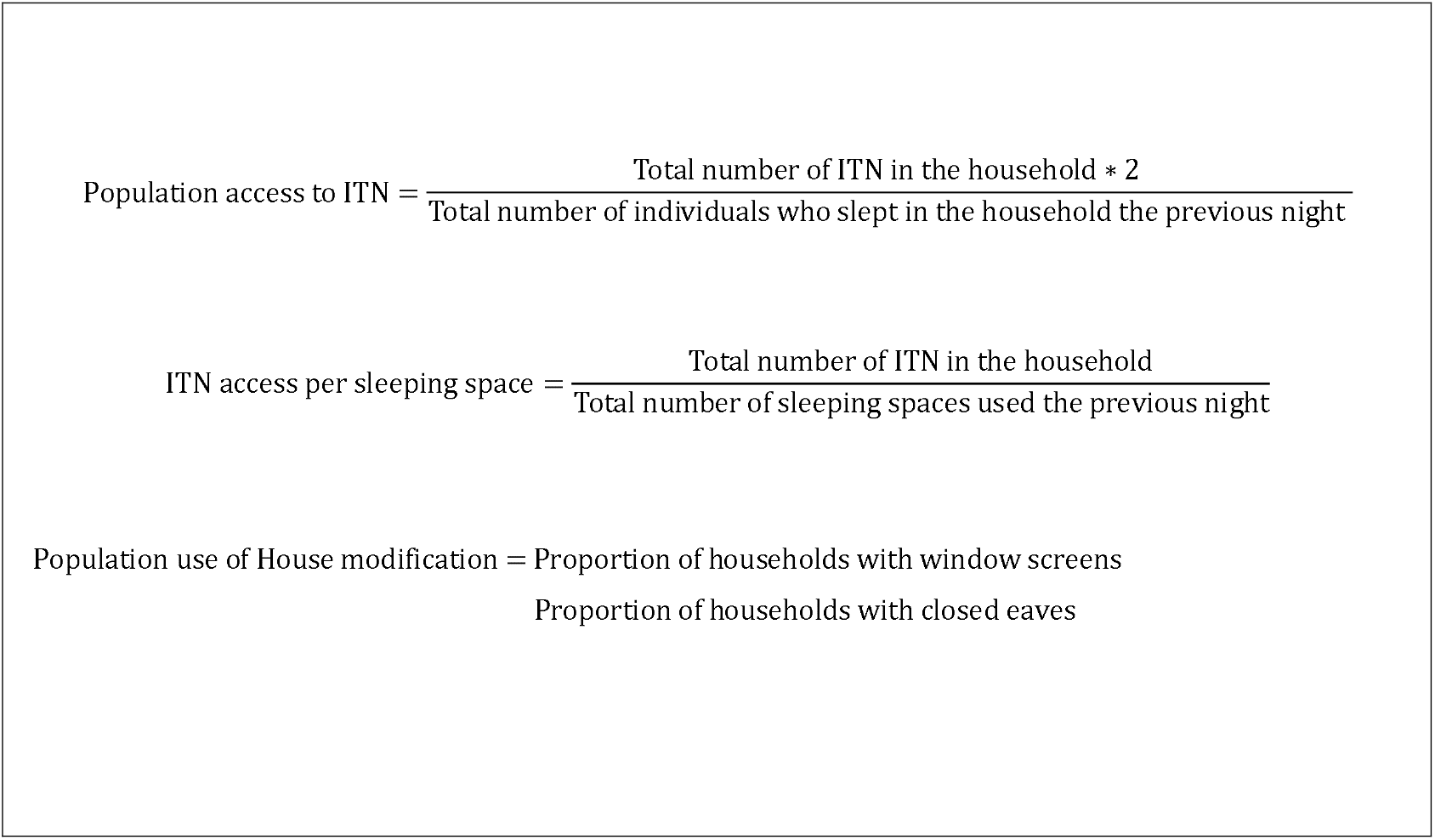
Indicators of ITNs access and the use of house modification.

### Estimating the association between mean distance of households to breeding sites and vector control

The geographical distance between the household and nearest mosquito breeding sites was measured using ArcGIS 10.3 [39]. The geographical locations of permanent and temporary breeding sites of all *Anopheline* species during the wet season in four villages of Bagamoyo district were collected with handheld GPS (etrex 10), and confirmation of larval presence in each breeding sites was done by larval dipping (Adam Saddler pers. comm). Shapefiles for the Tanzania districts were obtained from the National Bureau of Statistics (NBS) website and the Ruvu river shapefile was extracted from OpenStreetMap [40]. The mean distance of each surveyed households to breeding sites was categorized into two levels: ≤ 609.5 metres and > 609.5 metres, maintaining the mean travel distance of culex [41], as it is noted that most of the breeding sites contain *Culex quinquefasciatus* [42].

## RESULTS

### Demographic and socioeconomic characteristics of households

A total of 7,240 households in Bagamoyo and 1,241 in Ulanga districts were surveyed and their demographic characteristics detailed [Additional file 1]. Roughly 28% [95% CI 27 – 30] of households in Bagamoyo and 22% [16 – 29] in Ulanga consisted of more than five members. From data only collected in Bagamoyo, fewer households (9% [8 – 10]) were headed by young adults (18 to 24 years). There were more male headed households than female (24% [21 – 27]) and most of the respondents had primary education (71% [66 – 76]). The proportion of houses with more mosquito entries (windows and doors) were greater in Bagamoyo (44% [39 – 49]) than Ulanga (26% [18 – 37]), indicating more larger houses and wealthy households in Bagamoyo (23% [18 – 28]) than Ulanga (7% [5 – 11]). About 46% [6 - 91] of the houses in the population were located within a mean distance of 609.5 metres to a mosquito breeding site (Table 1).

**Table 1:** Demographic, socioeconomic and house characteristics

| **DISTRICTS** |  | **BAGAMOYO** | **ULANGA** |
| --- | --- | --- | --- |
| **Variables** |  | **Household Characteristics n (%)** |  |
| Total (N) |  | **7,240** | **1,241** |
| **Household size:** 1 to 2 |  | 1,804 (25) | 447 (36) |
| 3 to 5 |  | 3,373 (47) | 525 (42) |
| 6 to 20 |  | 2,063 (28) | 269 (22) |
| **Age group:** | 18 - 24 | 628 (9) |  |
|  | 25 - 49 | 4,192 (58) |  |
|  | 50 - 99 | 2,417 (33) |  |
| **Gender:** | Male | 5,533 (76) |  |
|  | Female | 1,707 (24) |  |
| **Education:** | None | 1,280 (18) |  |
|  | Primary | 5,158 (71) |  |
|  | Secondary or Above | 767 (11) |  |
| **House entry (windows and doors):** 1 & 2 |  | 1,460 (22) | 546 (44) |
| 3 & 4 |  | 2,301 (34) | 367 (30) |
| 5 & above |  | 2,998 (44) | 327 (26) |
| **Household location to breeding sites:** ≤ 609.5 metres |  | 864 (12) |  |
| > 609.5 metres |  | 741 (10) |  |
| **Wealth Quintiles:** Poorest |  | 1,211 (17) | 485 (39) |
| Poor |  | 1,361 (19) | 324 (26) |
| Middle |  | 1,531 (21) | 172 (14) |
| Rich |  | 1,486 (21) | 167 (13) |
| Richest |  | 1,651 (23) | 93 (7) |

### ITNs access in the population

Population access to ITNs (mean of households with an ITN for 2 potential users that stayed in the house the previous night) was estimated as 64% [95% CI 56 – 71]. ITNs per sleeping spaces (mean of households with enough ITNs to cover all sleeping spaces used the previous night) was also estimated as 63% [54 – 72] in Bagamoyo after 3 years of mass campaign. Both lower than the 80% coverage target of the Tanzania National Malaria Strategic Plan except in Ulanga where population access to ITN was 84% [78 – 91] and ITN per sleeping spaces was 93% [89 – 97] after 1 year of mass campaign. Differences between ITN access in the two locations are related to the timing of the survey.

### Factors associated with ITNs coverage in Bagamoyo and Ulanga

Univariate analysis revealed that on access to ITN was on average lower among households with fewer people and which head of house was younger, male, had little formal education, and poor, as well as in houses that had fewer number of windows and doors, and far from a breeding site in Bagamoyo and only lower among households with fewer people in Ulanga. While in multivariable analysis, all factors estimated were not significantly associated with access to ITNs except household size (Table 2). The consistent determinant of access to ITN was larger household size.

**Table 2:**
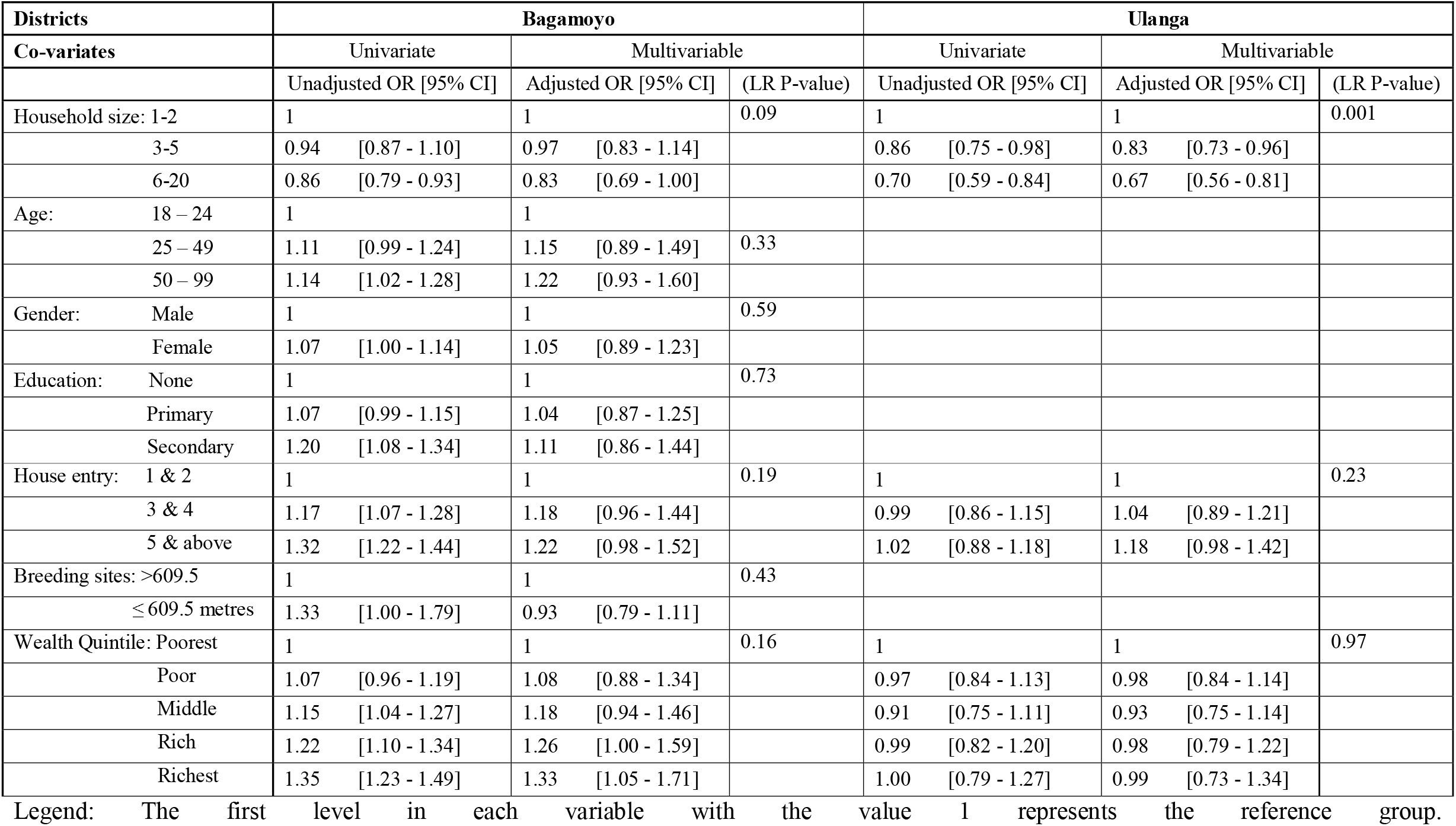
Factors associated with access to ITNs.

Similar to access to ITNs, on average enough ITNs per sleeping spaces was less common in Bagamoyo among poor households whose head had no formal education, as well as houses with fewer numbers of doors and windows located above 609.5 metres from a breeding site in the univariate analysis. However, there were no significant differences among the demographic and socioeconomic factors regarding ITNs per sleeping spaces in both Bagamoyo and Ulanga in the multivariable analysis (Table 3). Most households had even distribution of ITNs per sleeping spaces on average.

**Table 3:** Factors associated with enough ITNs to cover all sleeping spaces in the population

| Districts | Bagamoyo |  |  | Ulanga |  |  |
| --- | --- | --- | --- | --- | --- | --- |
| Co-variates | Univariate | Multivariable |  | Univariate | Multivariable |  |
|  | Unadjusted OR [95% CI] | Adjusted OR [95% CI] | (LR P-value) | Unadjusted OR [95% CI] | Adjusted OR [95% CI] | (LR P-value) |
| Household size: 1-2 | 1 | 1 | 0.26 | 1 | 1 | 0.81 |
| 3-5 | 1.05 [0.98 - 1.13] | 1.15 [0.97 - 1.35] |  | 1.04 [0.91 - 1.18] | 1.04 [0.91 - 1.19] |  |
| 6-20 | 0.96 [0.88 - 1.04] | 1.08 [0.89 - 1.31] |  | 1.00 [0.85 - 1.17] | 1.02 [0.86 - 1.20] |  |
| Age: 18 – 24 | 1 | 1 | 0.60 |  |  |  |
| 25 – 49 | 1.09 [0.98 - 1.21] | 1.14 [0.87 - 1.48] |  |  |  |  |
| 50 – 99 | 1.01 [0.90 - 1.13] | 1.10 [0.84 - 1.46] |  |  |  |  |
| Gender: Male | 1 | 1 | 0.57 |  |  |  |
| Female | 1.01 [0.94 - 1.08] | 1.05 [0.89 - 1.23] |  |  |  |  |
| Education: None | 1 | 1 | 0.57 |  |  |  |
| Primary | 1.13 [1.04 - 1.22] | 1.06 [0.89 - 1.28] |  |  |  |  |
| Secondary | 1.27 [1.13 - 1.42] | 1.15 [0.89 - 1.50] |  |  |  |  |
| House entry: 1 & 2 | 1 | 1 | 0.43 | 1 | 1 | 0.96 |
| 3 & 4 | 1.17 [1.07 - 1.27] | 1.14 [0.92 - 1.39] |  | 0.99 [0.86 - 1.13] | 0.99 [0.86 - 1.14] |  |
| 5 & above | 1.20 [1.11 - 1.31] | 1.06 [0.85 - 1.32] |  | 0.97 [0.84 - 1.12] | 0.97 [0.81 - 1.17] |  |
| Breeding sites: >609.5 | 1 | 1 | 0.24 |  |  |  |
| ≤ 609.5 metres | 1.31 [1.15 - 1.49] | 0.90 [0.76 - 1.07] |  |  |  |  |
| Wealth Quintile: Poorest | 1 | 1 | 0.42 | 1 | 1 | 0.99 |
| Poor | 1.04 [0.93 - 1.15] | 1.05 [0.85 - 1.30] |  | 1.01 [0.87 - 1.17] | 1.01 [0.87 - 1.17] |  |
| Middle | 1.13 [1.03 - 1.25] | 1.15 [0.92 - 1.44] |  | 0.95 [0.79 - 1.14] | 0.96 [0.79 - 1.17] |  |
| Rich | 1.20 [1.09 - 1.33] | 1.19 [0.94 - 1.51] |  | 0.98 [0.82 - 1.18] | 0.99 [0.80 - 1.22] |  |
| Richest | 1.35 [1.23 - 1.48] | 1.24 [0.97 - 1.58] |  | 1.00 [0.79 - 1.25] | 0.99 [0.74 - 1.32] |  |

2 Legend: The first level in each variable with the value 1 represents the reference group.

Association of other covariates access to ITNs and ITNs per sleeping spaces were not statistically significant in the univariate analysis [Additional file 2] and multivariable analysis [Additional file 3].

### Use of house modifications (window screens and closed eaves) in the population

Of the households visually observed for house modification, 61% [46 - 74] had window screens in Bagamoyo, while only 12% [8 - 17] had window screens in Ulanga. Some houses had closed eaves, 14% [10 - 20] of houses in Bagamoyo and 55% [45 - 64] in Ulanga. About half (49% [38 - 59]) of the sampled houses with screened windows in Bagamoyo also used an ITN and in Ulanga, 53% [44 - 63] of houses with closed eaves used an ITN. However, the proportion of houses in both districts with the combination of screened windows and closed eaves that reported use of an ITN the previous night of the surveys was less than 10%.

### Factors that are associated with the use of house modification in the population

In the univariate analysis, use of window screens was lower in poor houses in which head of household was older, female and had little education as well as houses with lower numbers of doors and windows close to a breeding site in Bagamoyo. Similar results were found in Ulanga, except that houses with fewer members were less likely to have a window screens. Of these findings, only household size, age and wealth of head of household, mosquito entry (higher numbers of doors and windows) and mean distance to breeding sites were strongly associated with the use window screens in both Bagamoyo and Ulanga in the multivariable analysis (Table 4).

**Table 4:**
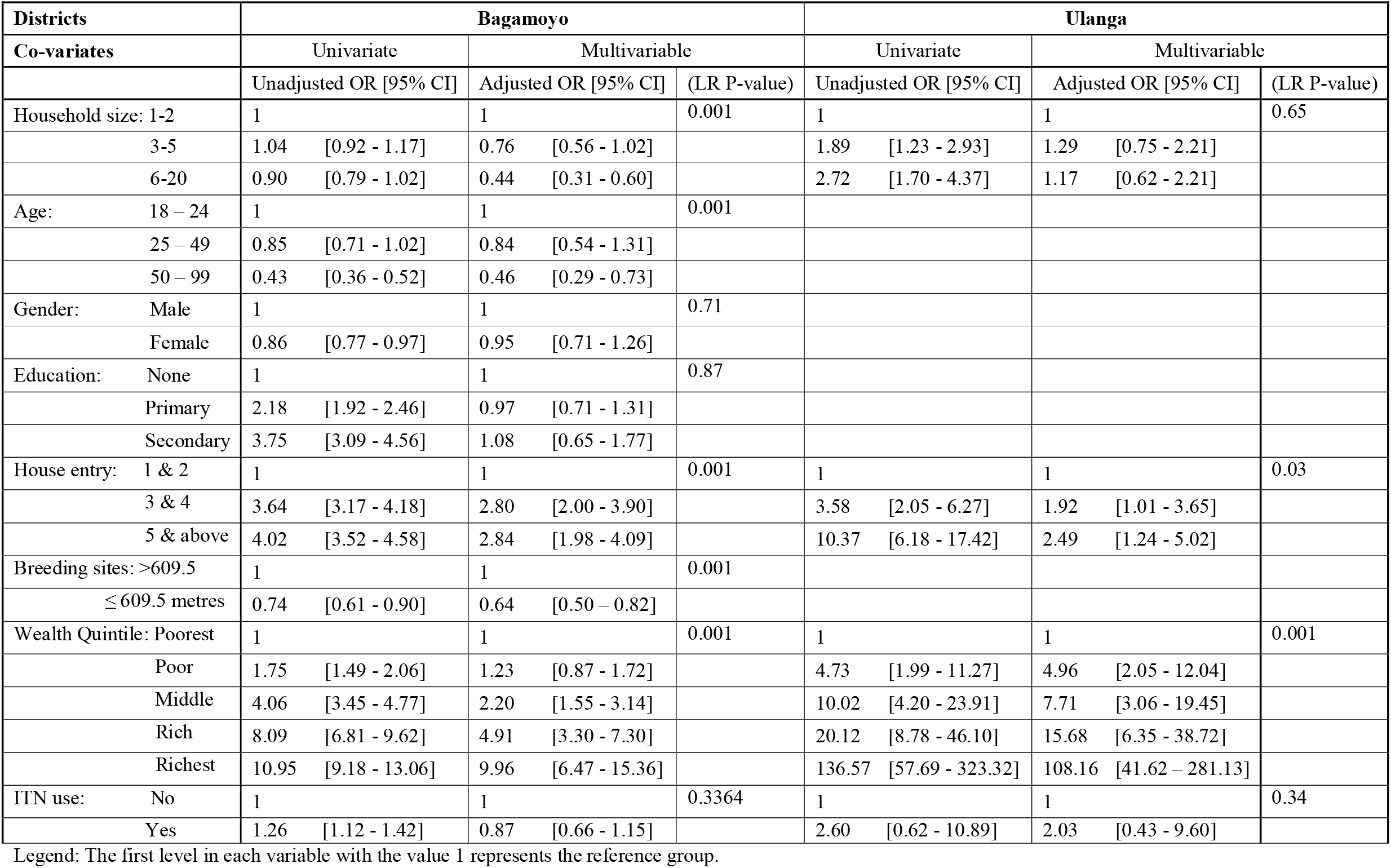
Factors associated with the use of window screens in the population

Larger households and those headed by older individuals, female and in the poor quintile were less likely to use closed eaves in Bagamoyo and similarly in Ulanga, except that households in the rich quintile had low usage of closed eaves in the univariate analysis. Although, only wealth and gender were significantly associated with the use of closed eaves, the proportion of houses with closed eaves was lower among poor households headed by make in Bagamoyo and higher in Ulanga as the poor households blocked their eaves with mud the multivariable analysis (Table 5). Other co-variates explored were not statistically associated with house modification in both univariate analysis [Additional file 2] and multivariable analysis [Additional file 3].

**Table 5:**
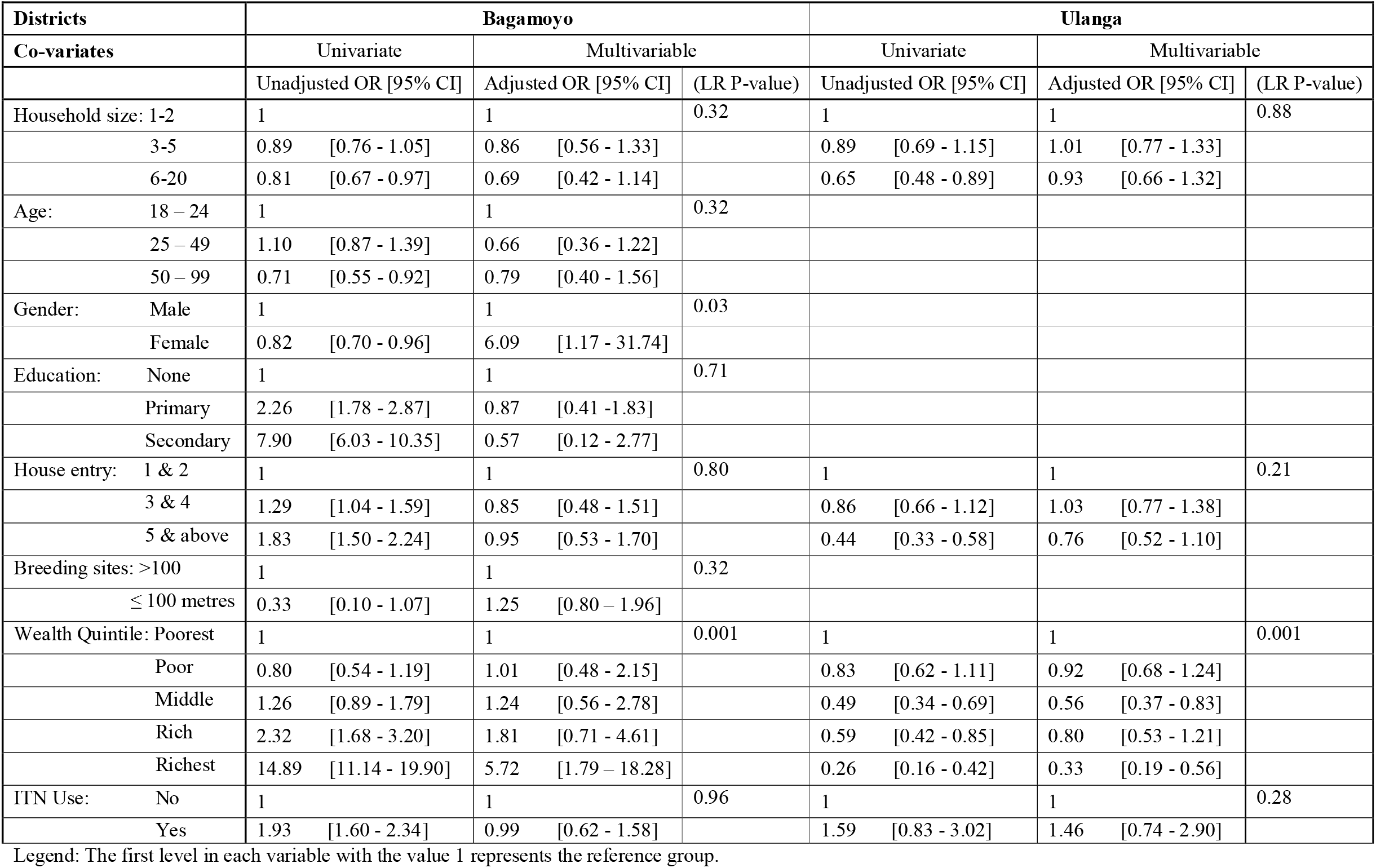
Factors associated with houses with closed eaves in the population

## DISCUSSION

### ITNs coverage in the population

Population access to ITNs after 3 years of mass distribution was lower than targets of the Tanzania National Malaria Strategic Plan. The difference between the access to ITNs in Bagamoyo and Ulanga (64% vs 84%) estimates that access declines at an average of 10% after the first year, this suggests that a minimum interval of two years ITN distribution campaigns is necessary to maintain high coverage of ITN, this is a concern as the average interval between campaigns in Tanzania is at least 4 years. Population access to ITNs is a more robust indicator of measuring ITN coverage [28] and was lower among large households even in Ulanga where the survey was conducted shortly after residents received ITNs from the national campaign. This finding has also been observed in the lake region of Tanzania where household numbers are on average larger, (Ikupa Akim pers. comm). Therefore our study also demonstrates that limiting the number of ITNs distribution by not necessarily taking into account individual household size in the population will continue to serve as hindrance to achieving high ITN coverage [28]. The Universal Coverage Campaign (UCC) implemented in Tanzania to cover all sleeping spaces [4] resulted in a higher access measured by ITNs per sleeping spaces (93%) rather than access to ITNs (84%) in Ulanga, this demonstrates that more than two people are using a single sleeping space. This is consistent as it was found that larger households had more children (< 5 years old), who were more likely to share sleeping space with their parent or a sleeping space occupied by more than two children. This has been found to decrease the durability of ITNs due to stretching of the net resulting in damage [43], [44]. Additional efforts are required to improve household access in order to maintain effective malaria control seen in previous years due to universal ITN coverage campaigns.

Family size is an important factor that needs to be considered when planning ITNs distribution campaigns. The last campaign in Tanzania achieved high coverage in Ulanga, considering the indicator of population access to ITNs, but it is necessary to ensure that the ITNs are actually where they are most needed, and in sufficient number in the larger households which are most likely to have the most malaria vulnerable groups (children under 5 and pregnant women). Mosquitoes are more attracted to households with an increased number of people [45], thus households with larger family size need to have adequate ITNs to impact malaria. ITNs distribution in schools may be a good idea for a keep up strategy [46]. School age children tend to have lowest access to ITNs [47], which emphasizes the utility of the school nets program in Tanzania. The slight influence of wealth was observed in Bagamoyo, which may suggest that rich people purchased ITNs when the ones they were given by the campaign became non-functional. Therefore, Behaviour Change Communication (BCC) strategy at the community level may be targeted towards all households to encourage care of ITN to last longer and continue the usage of damaged ITNs as against discarding while waiting for campaigns, as it has been shown that damaged nets remain insecticidal [48]. It is clear, however that more effort is needed in improving access to ITNs.

### House modification in the population

A possible explanation for the higher usage of window screens in Bagamoyo could be attributed to more economic development and availability of low cost screening as reported in a study conducted in Dar es Salaam – close proximity to study area [15], as compared to Ulanga, a remote rural area with low income and lack of access to window screening suppliers. Building materials are likely to be more expensive far from economic centres due to transport cost. The majority of the houses in Bagamoyo had opened eaves possibly due to low awareness of closed eaves as a tool to reduce indoor mosquito density [12] and high temperature during the daytime as found in neighbouring coastal Kenya (Mombasa) [49]. The use of open eaves is an efficient means of cooling houses [11] which is also a plausible explanation for the observed pattern, especially since Bagamoyo has an average temperature 2^°^C higher and greater humidity than Ulanga due to its proximity to the ocean. Wealthy households in Bagamoyo were more likely to have closed eaves where window screens were also in common use, while in Ulanga, the poorest households were more likely to close their eaves but were unable to afford window screens that gives protection from mosquitoes [50]. As window screening is clearly a function of wealth, it would be prudent for screens to receive a subsidy and for insecticide treated screens to be made more widely available as a long-term vector control intervention to protect individual users as well as the community – through killing vectors that contact the screens. The acceptance of closed eaves in the rural areas do not seem worrisome as it is already a common housing structure among modern houses owned by wealthy people in Tanzania [51]. However, in houses with indoor fires, mostly found in the remote areas, closing eaves is not advisable as may increase exposure to biomass particulate matter and induce respiratory illness [52]. There is evidence of high densities of indoor mosquitoes in houses with more entry points [16], which has led to the observed increased use of window screens in the population as the number of windows and doors increases.

Households headed by young individuals had more window screens installed. This group had more years of formal education and were wealthier than the older age group, as access to education in Tanzania has steadily improved over the last decades [53]. This might have made them more knowledgeable about the importance of the tool in preventing indoor mosquito entry and their wealth afforded them the opportunity to install them in their houses. Window screens are useful because they 1) require no compliance, 2) tend to be long-lasting and 3) protect all members of a household. This also serves as an indicator that younger people are adaptive to the uptake of new tools for controlling malaria. However, the use of window screens alone is not sufficient to control malaria [54]. The incorporation of insecticides into window screens would improve the efficacy of this tool against mosquitoes as it has improved the protective efficacy of conventional bednets and other vector control tools [55]. In addition, It is logical that maximising the concurrent use of multiple vector control tools, i.e. combination of ITNs and insecticidal house screening may assist in transiting the current malaria phase in Tanzania to elimination [56], [57] and may also protect against other vector-borne disease such as dengue [10].

The study area in Tanzania provides conducive breeding sites based on ecological conditions and human behaviour, hence high mosquito density notable for nuisance biting [58]. Our study found that window screens were less common among households at an average of 609.5 metres to a breeding site. This is because the poorest households were located at closer proximity to breeding sites and couldn’t afford the installation of window screens, which means that poorer households are exposed to higher risk of mosquito bites. It is reported in another study conducted in Tanzania that most of the breeding sites contain *Culex quinquefasciatus* which are notable for nuisance biting [42] and transmission of other human pathogens such as viruses and filarial worms [59]. This supports the essence of providing a subsidy for window screens to reduce the exposure of the poor households to vector-borne diseases.

### Limitations

Our study was affected by non-response bias as shown in Figure 3 and desirability bias, that might have influenced ITNs access estimates [60]. It was suspected that respondents denied owning at least one ITN in the household due to an expectation of compensation in the form of ITNs after the study. Therefore, it was necessary to use number of ITNs reported to be used the previous night as a proxy for ownership, it was more adequately measured as interviewers asked the use of each ITN in respect to where it was used in the household. Ideally, surveys of ITN access should visually inspect the presence of nets in households whenever possible to reduce the possibility of respondents giving misleading answers. The study was unable to establish a detailed comparison between the study areas because the age, sex and educational status of head of households as well as the geocoordinates of the breeding sites in Ulanga district were not collected. Nevertheless, the study recruited a very large number of participants which gave the study strong power to correctly establish association between the outcomes and covariates. House structures which included window screens and closed eaves were visually recorded by interviewers, and thus accurately observed.

## CONCLUSION

This study demonstrates the need to improve access to ITNs especially among larger households in the population, due to the fact that access already dropped within 3 years of distribution and among larger family even just after recent government distribution. Our study suggests that large family households should be specially targeted in keep-up and universal coverage campaigns. It is evident in the study that the wider use of window screens could largely be improved by government subsidies. Therefore, it is necessary to put strategies in place to improve access to window screens widely through low cost screens and insecticidal screens to maximise the use of the combination of ITNs and insecticidal house screening to further control malaria as well as other vector-borne diseases.

## Data Availability

All data are available and archived in the Ifakara Health Institute (IHI), Bagamoyo branch.

## Abbreviations

ANC: Antenatal Care
BCC: Behavior Change Communication
DHS: Demographic Health Survey (Tanzania)
EPI: Expanded Programme on Immunization
GIS: Geographical Information System
IHI: Ifakara Health Institute
IRS: Indoor Residual Spray
ITNs: Insecticide Treated Nets
NMCP: National Malaria Control Plan
NMSP: National Malaria Strategic Plan
PCA: Principal Component Analysis
SES: Socioeconomic Status
UCC: Universal Coverage Campaign
WHO: World Health Organization

## DECLARATION

### Author’s contribution

OGO conducted study analysis and paper writing. AR contributed to the statistical analysis and contributed to manuscript revision. YPM contributed to the major revision of the manuscript. OJ contributed to the manuscript’s revision. SM contributed to data management and paper writing. DFM assisted substantially with data management and involved in manuscript writing. SJM conceived and designed the study, also contributed to analysis and paper writing. All authors contributed to the final manuscript revisions and approved the final version.

### Funding

The study was funded by the University of NOTRE DAME (UND Fund #261655).

#### Acknowledgement

We acknowledge the efforts of the research team (technicians) of the Ifakara Health Institute, Bagamoyo (IHI) and participants for taking their time to collect and provide data, respectively. Dr. Adam Saddler provided advice and data source on the breeding sites, PD Dr. Christian Schindler contribution on statistical analysis was helpful, as well as Raphael S. von Büren who provided guidance on proximity analysis on ArcGIS. Dr. Albert Kilian and Dr. Hannah Koenker provided guidance on calculation of population access to ITNs.

### Availability of data and materials

Not applicable.

### Consent for publication

Not applicable.

### Ethics approval and consent to participate

Ethical clearance to conduct the survey was obtained from the Medical Research Coordinating Committee of the National Institute of Medical Research NIMR/HQ/R.8a/Vol. IX/1899 and the Institutional Review Board of the Ifakara Health Institute IHI/IRB/EXT/18 - 2015 and IHI/IRB/EXT/15 - 2017 and NIMR/HQ/R8.a/Vol IX/2270.

## Additional files

Additional file 1: Distribution of demographic and socioeconomic characteristics, and ITNs coverage and use of house modification.

Additional file 2: Univariate analysis of the factors associated with ITNs coverage and use of house modification.

Additional file 3: Multivariable analysis of the factors associated with ITNs coverage and use of house modification.

